# Autism Classification using Visual and Behavioral Data

**DOI:** 10.1101/2022.10.28.22281655

**Authors:** Nafisa Sadaf Hriti, Karishma Shaer, Farhan M Nafis Momin, Hasan Mahmud, Md. Kamrul Hasan

## Abstract

Currently Autism Spectrum Disorder (ASD) is diagnosed via the combination of multiple medical tools and screening tests that require extensive amounts of time and money. Autism diagnosis can be formulated as a typical machine learning classification problem between ASD patients and a control group consisting of neurotypical individuals. In order for this to yield accurate results, large datasets with different modalities are required. However, the unavailability of such robust datasets stands as a threat to this automated diagnosis. To resolve this, we propose a method of Autism Classification using Visual and Behavioral Data. The proposed technique relates datasets of two modalities (visual and behavioral) collected from similar participants by generating common attributes among the records and distributing these records into sub classes. Then records within these subclasses are combined to form an integrated dataset. Finally, decision level fusion is performed on the multimodal data. The main contribution of our work can be outlined as follows: an accuracy of 97.57% in autism classification has been obtained from the integrated data, which is higher than detection from only visual data, we have shown that combining data within sub classes based on common attributes is more accurate than combining them arbitrarily, and finally, we have introduced a novel, integrated multimodal dataset in the ASD domain.

## 3 Introduction

Autism Spectrum Disorder (ASD) is a developmental disability that can cause significant social, communication and behavioral challenges [1]. There are many sub-types of ASD that occur due to biological and environmental markers. As the name Autism Spectrum Disorder suggests, this disability can manifest in a set of symptoms that is unique to each patient. It can range from the patient being highly intelligent to being severely challenged. Some of the clinical features that are characteristic of autism are sensory processing disorders, gastrointestinal disorders or seizures. Furthermore, ASD can give rise to other issues such as poor sleep quality, difficulty in paying attention, anxiety and depression.

There is no singularly definitive medical test, like an MRI scan or a blood analysis etc, to diagnose the disorder properly. In the current procedure for diagnosis, medical professionals examine the patient’s history of development and observe their behavior to reach a decision. This is a completely manual process that requires vast experience and extensive knowledge of psychology and neuroscience.

Past research has shown that ASD can often be detected at early ages - 1.5 years or younger. By the age of 2, diagnosis by a medical expert can be considered definitive. Despite that, for many children the disorder is identified at a much later age. Machine learning is found to be a useful technique for the early diagnosis of ASD [2].Such long delays prevent children with ASD from getting the proper help required on time. Treatment can begin earlier if children can get diagnosed early. Applied Behaviour Analysis, or ABA [3], is the application of the science of behaviour analysis that helps those on the autism spectrum to lead fulfilled lives[4]. Early intervention programs, particularly for children, have been shown to yield significant benefits in academic achievements, social behavior, educational progression and attainment, while also improved labor market success.

Machine learning algorithms can be utilised to classify the presence or absence of ASD in a random child when characteristic features are available. By using a large Personal Characteristic Data (PCD) dataset, nine automated machine learning models had been developed and compared [5]. However there was presence of heterogeneity in the dataset and the small size of the dataset affected the generalization performances of the models. Experiments have been conducted to perform video gesture analysis for ASD Detection [6] by recording a set of video clips of reach-to-grasp actions performed by children with ASD and IQ-matched normal children. A dataset on eye movement of children with ASD Disorder [7] had been designed and more recently Gerry [8] built an image dataset of ASD and non-ASD children; he extracted and used the facial features from the images of the dataset and fed them into a classifier. Emotional and behavioral information [9] and brain imaging markers[10] have been used in identifying the likelihood of autism in children as well. There are multiple small datasets on ASD containing particular data types. Deep neural network (DNN) have been used in recent years to identify patients with ASD using QCHAT datasets [11]. Simple model architectures benefit from the availability of a large number of data points since outliers are easier to classify, X-fold cross validation can be used and the underlying distribution of the data is clearer.

To our knowledge, there is no existing work done for multiple ASD dataset integration. Our goal is to integrate multiple datasets of different modalities into one efficiently large dataset with higher quality that can be used in further research work on diagnosis tools and high accuracy ASD screening. The efficiency of any diagnostic classifier of ASD heavily depends on the size and diversity of the data available. We believe the existence of such an enriched dataset can enhance the accuracy of ASD classifiers.

## 4 Materials and Methods

### 4.1 Background Study and Literature Review

#### 4.1.1 Classification of Autism Spectrum Disorder (ASD)

Currently, the manual diagnosis requires direct interaction with medical professionals to assess whether a child has ASD or not. It requires a child’s developmental history, responsiveness, behaviour, attention span, and intelligence to be checked. By the age of three, children with ASD display clear symptoms such as sensory problems, difficulty in communication, poor coordination, as well as declining emotional and social well-being.

Investigations into the facial structure of children have revealed insights into visual features such as increased facial masculinity in the ASD group relative to control group [12]. A recent study reveals that patients have a common pattern of distinct facial deformities, which can be used by a deep learning model (MobileNet and two dense layers) for classification [13]. Much of the previous work on ASD diagnosis used ideal clean images front-facing, obstruction-free, little variety in expressions of the subjects and clear backdrops. In this study, contrary to what was done before, the facial photos had been obtained from real scenarios online and the aggregate of these images produced the dataset. In our experiments with image data, we have used the model and image dataset originally used in this paper. Behavioral data of 1054 records[14] had been used to classify ASD with high accuracy in a recent work[15].

#### 4.1.2 ASD Datasets

In [6], researchers recorded a set of video clips of reach-to-grasp actions performed by children with ASD and IQ-matched typically developing (TD) children and made a video gesture dataset, consisting of 40 subjects (ASD: 20). [7] contains a dataset of eye movements of children with ASD disorder, which consists of 300 natural scene images and the corresponding eye movement data collected from 28 children (ASD: 14). Autism Brain Imaging Data Exchange (ABIDE) [16] has aggregated functional and structural brain imaging data. ABIDE I contains resting state functional magnetic resonance imaging (R-fMRI), anatomical and phenotypic datasets and ABIDE II consists of over 1000 additional datasets with greater phenotypic characterization, particularly regarding associated symptoms of ASD. The autism adult screening dataset in [17] contains 704 instances (ASD: 189). The dataset in [14] consists of behavioral data of 1054 instances with 18 attributes. It was used in our experiments and in [15]. The dataset in [8] was also used in our experiments and in [13], and contains facial images of children with 3014 instances (ASD: 1507).

A study in 2020 [13] had very promising preliminary results with an accuracy of 94.6%, but the image dataset used had duplicate images, improper age ranges, and lack of validation about the conditions of the individuals in each photo. According to this work, improving the dataset could yield better results, which we have attempted to execute in our experiment. Sufficiently large datasets on ASD are not easily available, and even if they are sizable, they mostly contain data of singular modality, for example only images, or only Personal Characteristic Data (PCD), or video, or hand gestures etc. The two datasets that we found most promising for our experiment have been collected from subjects that are similar in age group, gender and ethnic background. Previous work done in the field of ASD has proven that integrated features can improve Autism detection [5]. Successful integration of PCD and rs-fMRI taken from the same subjects has also shown excellent results [18]. Lastly, an integrated model using both Visual and Behavioral Data for diagnosis has not been implemented in the domain of autism.

#### 4.1.3 Gender and Race Prediction

Gender and race predictions of satisfactory accuracy can be obtained using classification algorithms when organic real-world visual data is input. Large refinement in this research area has occurred due to its large scale real-life application. Convolutional Neural Networks (CNNs) have been a breakthrough in this field and their usage in classification has boomed due to their notable improvement in results. Four widely used face detection tools, which are Face++, IBM Bluemix Visual Recognition, AWS Rekognition, and Microsoft Azure Face API were evaluated using multiple datasets to determine their accuracy in inferring user attributes, including gender, race, and age [19].

The tools mentioned provide predictions of age and gender, along with ethnicity in some cases. The classes pre-specified for ethnicity classification are white, black, or Asian- these classes divide the ethnicities in broad strokes and a more accurate label for them is “race”. Existence of multiple ethnicities within a race is possible, like the ethnicities “Korean” and “Japanese” may be contained within the race, “Asian.”

The dataset that we found to be most promising for our work is the UTK Dataset, a large-scale face dataset that was used in [20]. It is composed of over 20,000 images, each containing relevant age, gender and race labels. The age ranges from 0 to 116 years. Compared to the FairFace dataset of [21], UTK Dataset has a higher ratio of images of toddlers to adults. The ethnicities are as follows: White (42.5%), Black (19.1%), Asian (14.5%), Indian (16.8%) and Others (7.14%). As is evident, similar to our visual and behavioral datasets, the UTK Dataset is also white dominant, thus making it consistent with our work. It has an almost equal distribution of gender with males comprising 52.3% and females 47.7% of the entire data.

#### 4.1.4 Data Integration

According to [22], modern data generated in many fields are in a strong need of integrative machine learning models in order to better make use of heterogeneous information in decision making and knowledge discovery. For successful and robust analysis of data, congregating data from multiple sources before creating a generalised learning model on that data is a fundamental practice.

There are three categories of data fusion techniques: early, intermediate, and late integration. In early integration methods, all features are concatenated into a vector and then fitted onto an unsupervised or a supervised model; in late integration methods, separate models are first learned using their corresponding feature subsets, and then their outputs are further combined to make the final determination[22].

A comparative study [23] considered only decision level fusion techniques and observed that the performance of the highest confidence (HC) algorithm is the best among all the algorithms taken into account. It had an accuracy of 95.08% rate. The study revealed that arithmetic rules provide greater accuracy after fusion compared to abstract schemes or fusion based on rank.

Deep neural networks can be used for multi-modal learning and it can be extended to data integration too. Deep learning models of many types can be used on individual data sources. Information from the smaller networks can be combined in an integrative network. The deep neural network based multi-modal structure can merge the output of each independent sub-network as the network gets deeper.

A multimodal imaging study involved low-functioning young ASD patients of age 3–6 years. Classification performance reached an accuracy 88.8% and some prominent features were found. Machine learning-based analysis of MRI data was useful in classification of ASD patients from typically developing ones. Combination of two types of data had improved classification accuracy about 10% [24].

A co-clustering strategy proposed in [25], involves finding row and column clusters of a data matrix simultaneously and has been proven to be beneficial in high-dimensional settings in uncovering meaningful and explicable row groups even among high number of variables. It accounts for capturing similarities and differences among variables, as well as correlations between repeated measures among heterogeneous subjects.

### 4.2 Proposed Approach

#### 4.2.1 System Architecture

The two datasets consisting of the visual and behavioral data were obtained from children of the same age group since the symptoms of autism express themselves similarly depending on age. Both datasets had been filtered to keep children in the age range of 0 to 4 years. Initially common data clusters are identified between the two classes (autistic and non autistic) by filtering the records that have the same race and gender in both datasets. The clusters created were thus based on three attributes: race, gender and class label. The records corresponding to each cluster are merged to obtain the integrated dataset. The dataset is then trained through two binary classifiers corresponding to each modality of the data to generate class scores. These class scores are then combined using three different methods and the resulting scores of binary classification are evaluated.

**Figure 1:**
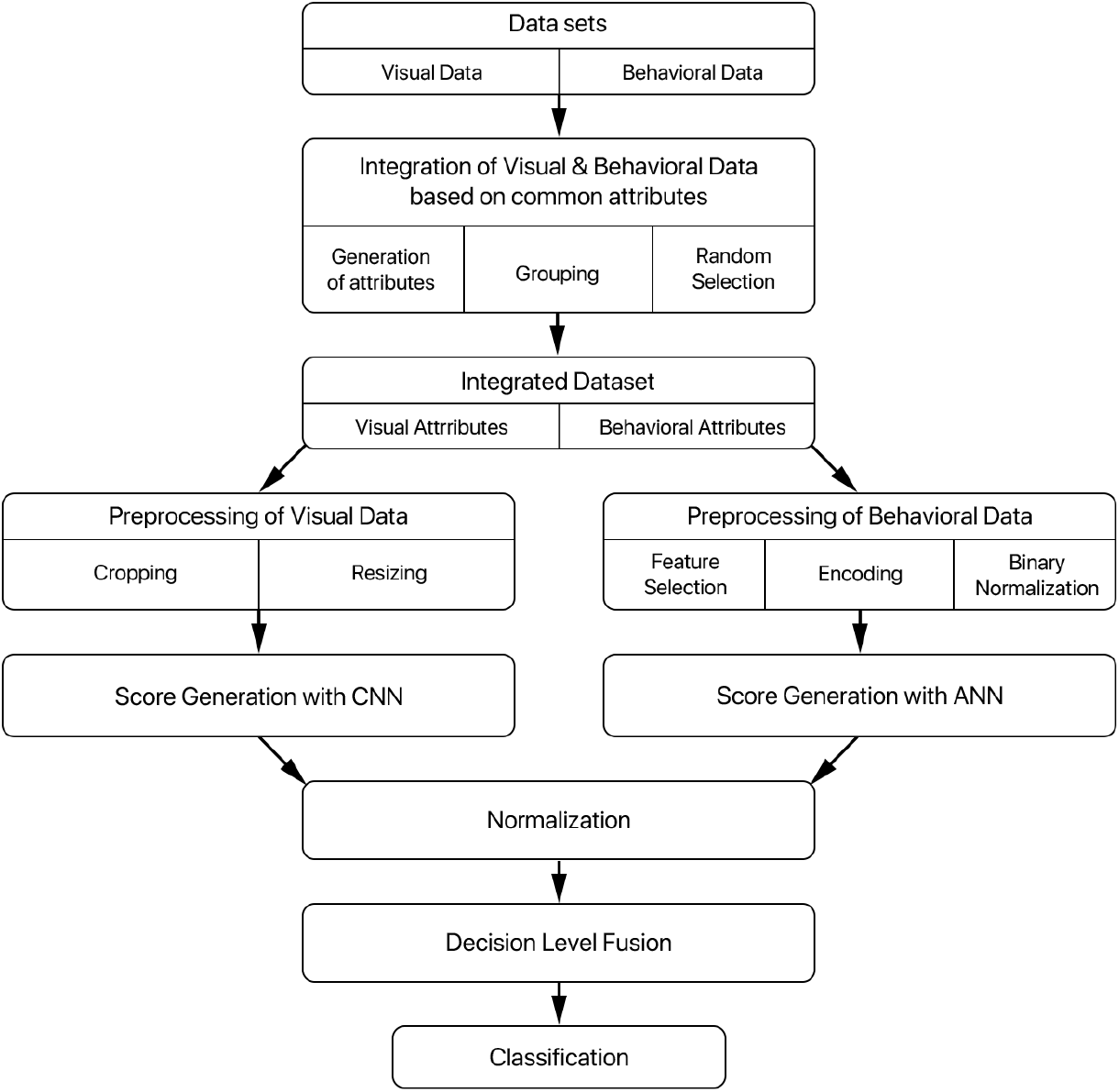
Proposed Approach.

#### 4.2.2 Integrated Record Generation

The image dataset being used in the proposed approach was obtained from Kaggle, which consists of 3014 images of both autistic and non-autistic children, equally divided in each class. The behavioral dataset is related to autism screening of toddlers and contains influential features to be utilised for further analysis especially in determining autistic traits and improving the classification of ASD cases. The dataset has 1054 records, ten behavioural features (Q-Chat-10) and other individual characteristics that have proven to be effective in detecting the ASD cases in behaviour science.

**Figure 2:**
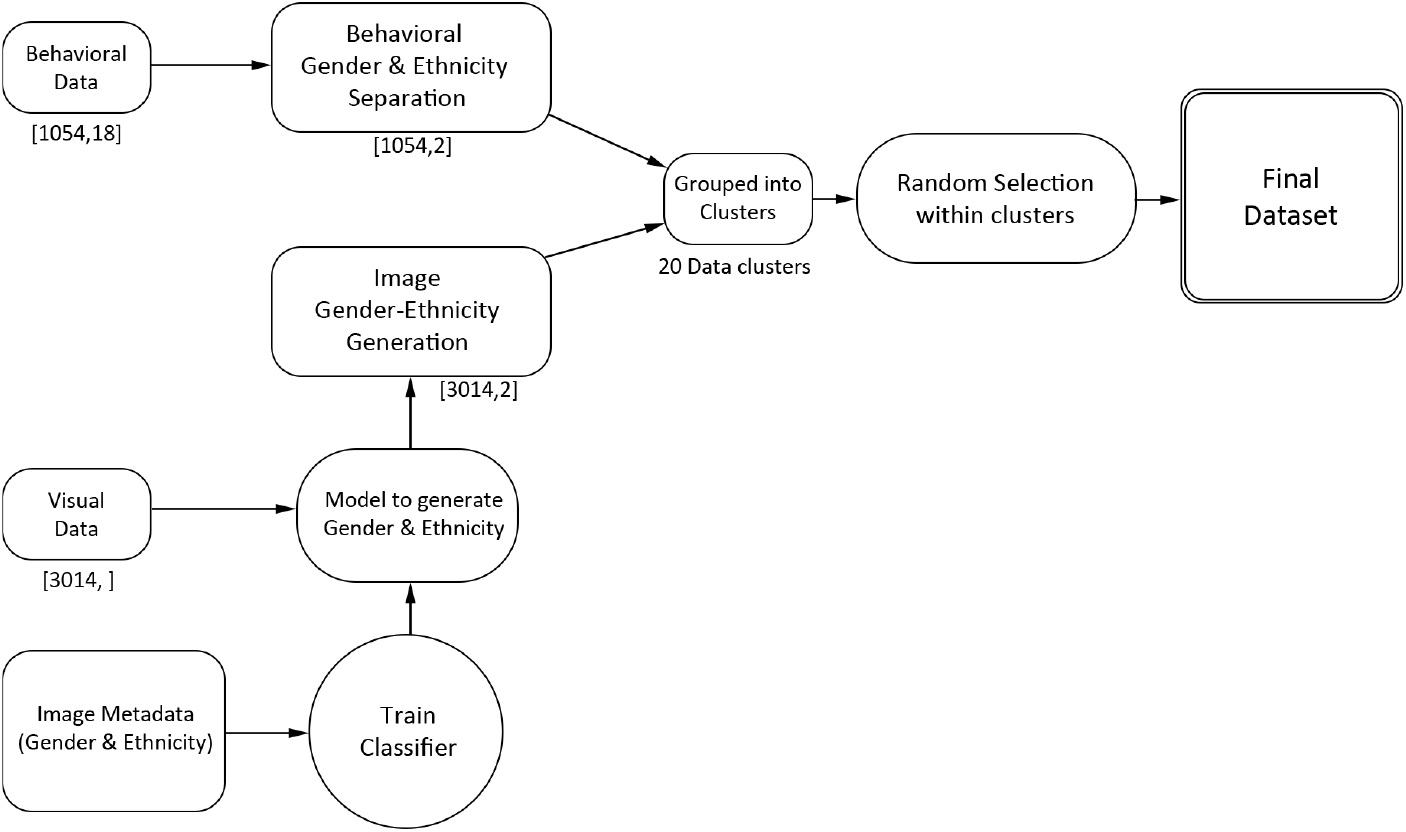
Integrated Dataset Generation.

Since the Visual and Behavioral Data being used have no common records and different modalities, two different methods of integration are required that approximate the relationship between the two datasets. Firstly, the relationship between the individual records of the two datasets are approximated. To find the association between two sets of data, predictive common attributes are generated using the visual data. The attributes that are generated are the Gender and the Race of the subjects in each of the facial images. Gender is classified into two classes: a) Male and b) Female. Race is classified into five classes: a) White, b) Black c) Asian, d) Indian, e) Others. In this process, a large-scale face dataset called the UTK Dataset [26] is used as our image metadat that consists of over 20,000 images, to train a multi-output CNN model that will output the predicted gender and race.

The multi-output CNN model consists of three branches: the age, gender and race branch respectively. Each branch follows a similar set of hidden layers: Conv2D with ReLu activation followed by BatchNormalization followed by Pooling followed by Dropout. This is then connected to two fully connected layers: the first with 128 nodes and the second has number of nodes equal to the number of classes (for instance, for gender branch there are 2 classes and for race branch there are 5 branches). The entire architecture of the multioutput CNN model is given below. After training, our visual data is fed into this model to obtain the predicted gender and race of each image. Apart from these two predictions, another attribute that is used to associate the records in the datasets is the binary class label: a) Autistic and b) Non Autistic. This procedure of visual metadata generation is depicted in the diagram below.

**Figure 3:**
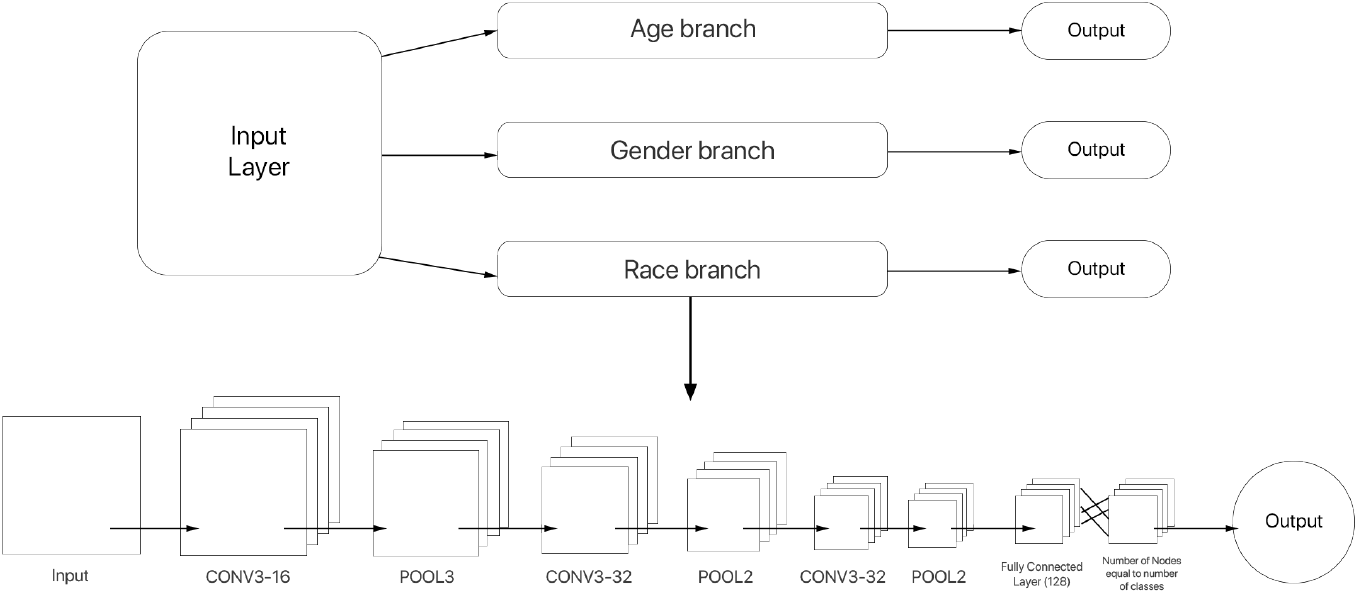
Multi-output CNN model Architecture.

**Figure 4:**
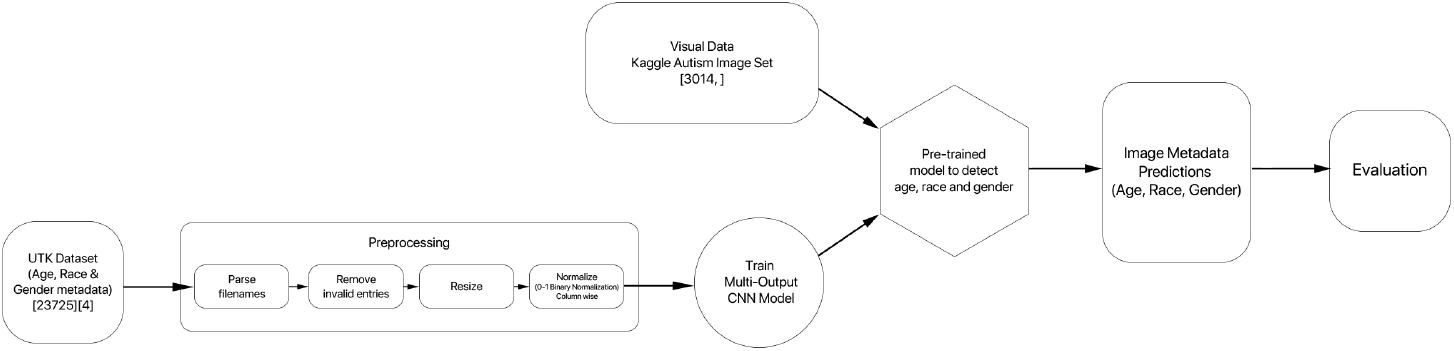
Generation of Visual metadata.

Taking all of these generated attributes into account, the whole image dataset is divided into 20 data clusters corresponding to each combination of gender, race and class label. Similarly, the tabular data of the behavioral dataset is also divided into 20 clusters. For each image in the visual dataset, a tabular record from within the data cluster is chosen arbitrarily and concatenated with the visual record to form a record in the integrated dataset. Each of the integrated records consists of an image and a corresponding tabular record of the behavioral data.

#### 4.2.3 Score Generation

The images and the behavioral data from the integrated records are trained on two different classifiers to generate score values. Decision level fusion of the datasets are done due to the multimodal nature of the data, which results in different preprocessing, feature generation and training to be best suited to each data type.

**Figure 5:**
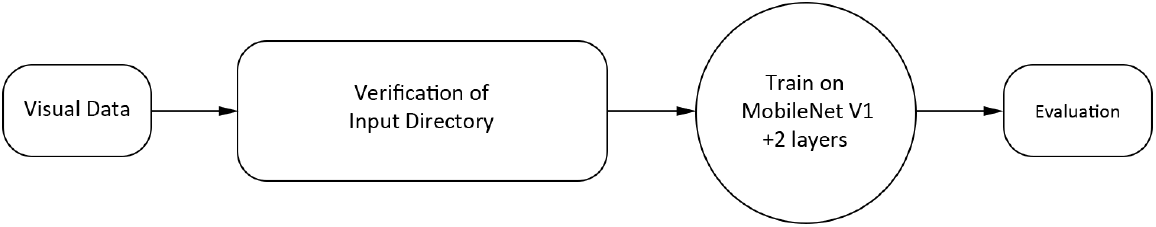
Processing of visual data.

Since the visual data is already cropped and cleaned to have uniform sized images, there is minimal preprocessing required on it, as shown in the figure below. The input directories of images are verified and after that the images from the integrated dataset are trained on a MobileNet classifier with two augmented dense layers. MobileNet has been shown to be just as accurate as the larger CNN architectures while significantly reducing the computing. To perform deep learning on the dataset, MobileNet is utilized followed by two dense layers. The first layer (with L2 regularization and ReLu activation is dedicated to distribution, and allows customisation of weights to input into the second dense layer. A dropout of 0.4 is applied to the first layer to prevent overfitting. Then the second dense layer allows for generation of the logit scores. The second layer consists of two nodes, with no activation function applied to it. These scores are the prediction weights of the of the classes given by the classifier. The architecture model of the CNN is given below.

**Figure 6:**
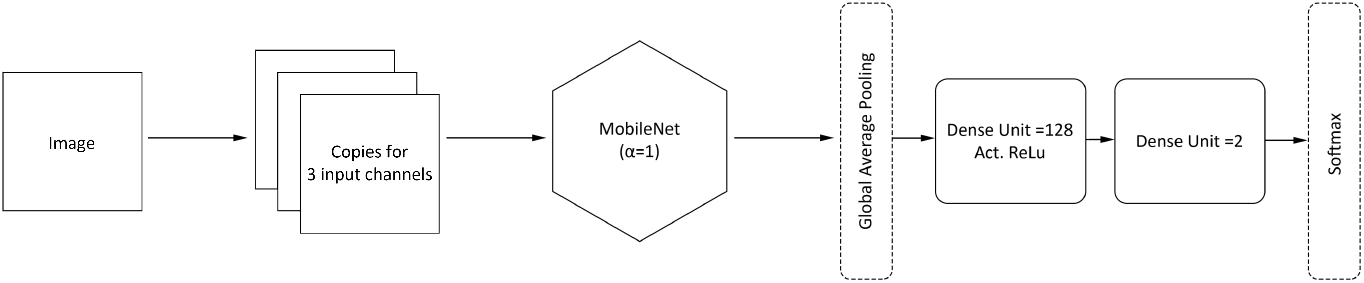
CNN model Architecture.

The behavioral data records are taken from the integrated dataset and trained separately. The data is preprocessed before being trained with a neural network to generate logit scores. Feature selection step is used to select relevant records from the dataset. The 10 attributes pertaining to the AQ-10 questionnaire were selected, along with the gender, race, age and whether any family member has ASD. After that, the data is cleaned to remove all the records with incomplete or missing information. Then the categoriacal data is encoded into numerical values to be used in the neural network. The whole numerical dataset is normalised in order to prevent any bias to be coming from the varying range of the attribute values. The behavioral data is then passed through an Artificial Neural Network (ANN) consisting of 3 hidden layers: with 64, 128 and 64 layers. A dropout rate of 0.8 is applied to prevent overfitting. The last layer of the ANN consists of two nodes without any activation function applied to them. These two nodes generate two score values that are used to calculate the final prediction of the combined model. The architecture model of the ANN is given below.

**Figure 7:**
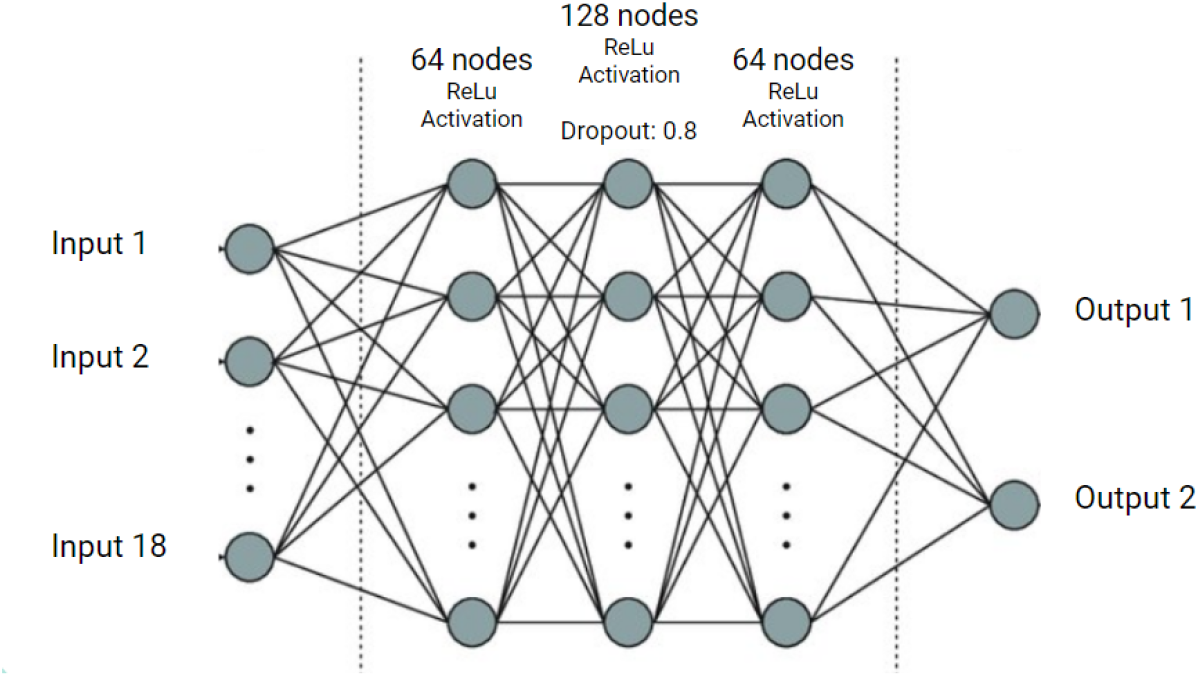
ANN model Architecture.

The figures below illustrate the processing on behavioral data and the hyperparameter setting of both ANN and CNN models, respectively. For the ANN model, we chose the best set of hyperparameters (number of nodes = 128, dropout = 0.4, classifier = adam) which gave an accuracy of 100%. For the CNN model, we chose the set of hyperparameters as used in [13] (dropout = 0.4, regularization strength = 0.015) which gave an accuracy of 93.57%.

**Figure 8:**
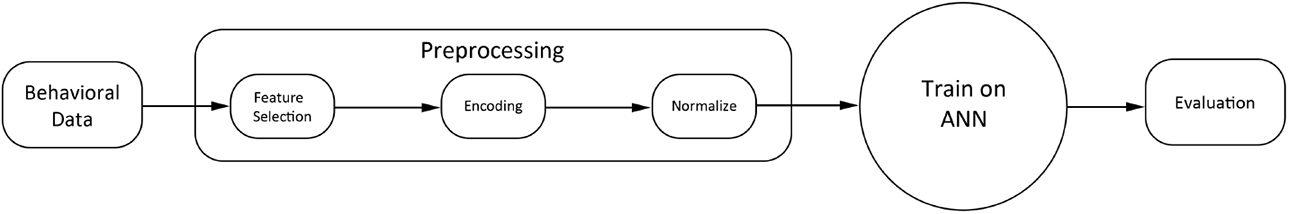
Processing on behavioral data.

**Figure 9:**
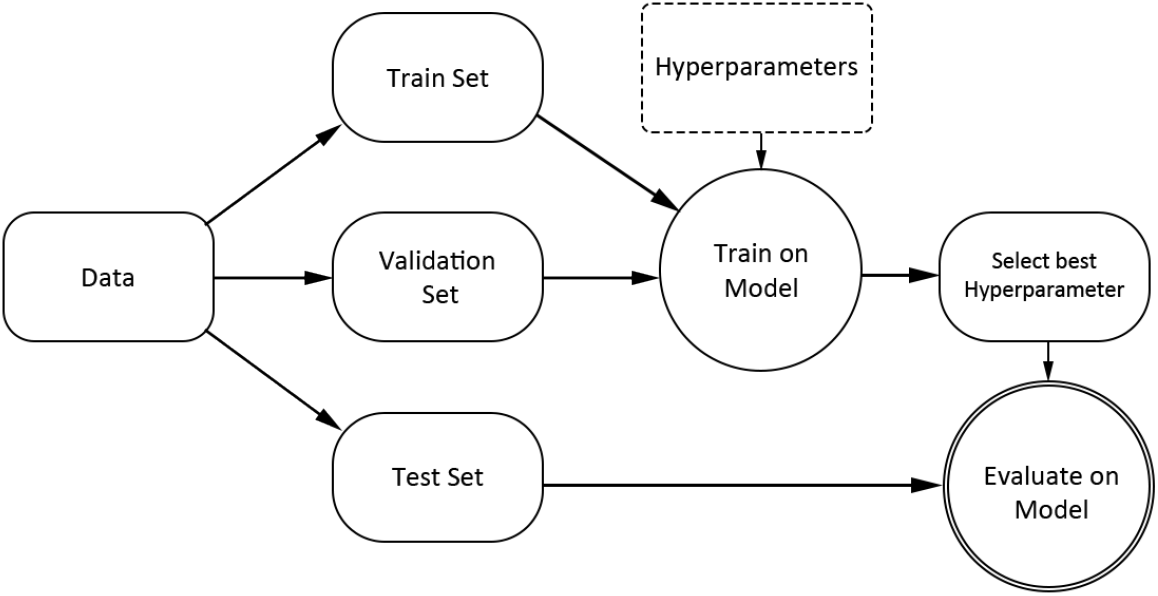
Hyperparameter setting.

#### 4.2.4 Decision Level Fusion

The scores obtained from the ANN and the MobileNet architecture are used to give final prediction of the class labels for the test records. There are two scores obtained from the ANN and two from the MobileNet architecture. This is to ensure that there is no bias of any dataset due to unequal number of scores from any dataset. The range of values can also cause bias of particular attributes, so all the scores are normalised. After that, to ensure that the best decision level integration method is used, three different methods are used to integrate the scores and the final predictions are evaluated.

**Figure 10:**
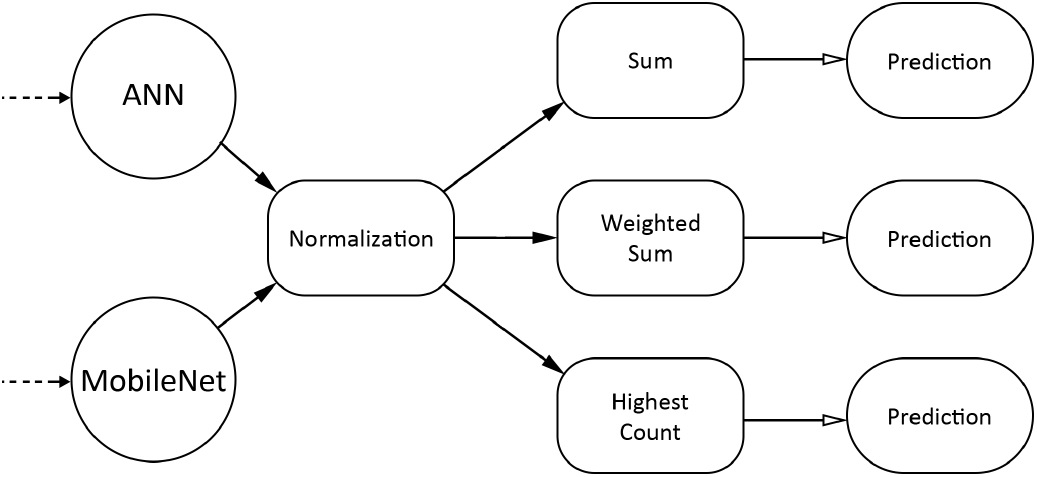
Decision Level Fusion.

The three methods are: a) Sum- The average of the scores are taken for each class by adding corresponding scores from each dataset and dividing by 2 (number of datasets). b) Weighted Sum- This is similar to sum, but before addition, a weight is assigned the scores from each dataset. c) Highest Count (Max)This method compares the scores from each dataset and takes the maximum value. After applying these methods to integrate the scores, each score is passed through a softmax classifier to generate the probability of the integrated record belonging to each class.

### 4.3 Experimental Setup

The proposed approach to detect ASD from two distinct datasets were tested on the Integrated records of the dataset and on each dataset individually. Large datasets will require exponentially more time to compute the Cartesian product of the records, so instead the image data rows are randomly concatenated with the behavioral data within each sub class. To test the hypothesis that Integrated Dataset would indeed perform better than initial datasets, three experiments are performed and the performance is evaluated in terms of sensitivity, specificity, accuracy, f-score, precision and recall each time. The experiments are: a) ASD Detection using the Visual Dataset, b) ASD detection using the Behavioral Dataset c) ASD detection on the Integrated Dataset. Three methods of decision level fusion are used, namely: sum, weighted sum and maximum. All three methods of fusion are compared with each other by evaluation based on the sensitivity, specificity, accuracy, f-score, precision and recall. Thereby we can experimentally discern if the Integration method is useful and if it is, which type of integration gives the best performance.

Our experiment utilised three datasets, Dataset 1 and Dataset 2 contained the behavioral and the visual data respectively to be integrated. Dataset 3 contained the gender and ethnicity labeled facial images that have been used to train the classifier that generated the image metadata before integration.

#### 4.3.1 Dataset Preparation

Each dataset underwent the necessary preprocessing steps that made them suitable to be used with the classifier models. The preprocessing required and the overview of each dataset have been outlined below.

##### Dataset 1 Description

Our data set 1 “Autistic Spectrum Disorder Screening Data for Toddlers “, which was also used in [15], was recorded via Q-Chat-10: Quantitative Checklist for Autism in Toddlers. It consists of 1,054 instances with each instance having 18 attributes (including the class labels). The attributes are of categorical, continuous and binary types. Q-Chat-10 gives scores based on ten behavioral attributes which it calculates from a questionnaire. For preprocessing, the features relevant to autism detection are selected, then the categorical features were encoded, and finally 0-1 normalization was performed on the values to ensure they are standardized within a fixed range.

##### Dataset 2 Description

Our data set 2 “Detect Autism from a facial image”, which was also used in [13], was obtained from Kaggle, and consists of 3,014 children’s facial images in total. The images are evenly split between two classes: autistic (1507) and non-autistic (1507). Each image is in 224 × 224 × 3 jpg format. The images were obtained from online, both through Facebook groups and through Google

Image searches. Once all the images were gathered, they were subsequently cropped so that the faces occupied the majority of the image. Prior to training, the images are split into three categories: train, validation, and test. This dataset has several versions and the ones we used in our experiments are version 1 and version 9 and compared the performances with both. The version 1 contains few duplicate images which could account for the slight increase in accuracy as mentioned in [13]. Since the images of this dataset had already been cropped and resized, the size of the images was compatible the model and no further preprocessing was necessary.

##### Dataset 3 Description

Our data set 3 “UTK Dataset” [26], which was also used in [20], is a largescale face dataset. It consists of over 20,000 images in total. The images cover large variation in pose, facial expression, illumination, occlusion, resolution, etc. The images are evenly split between two genders. Each image has its respective annotations of age, gender and race. Each record is stored in the following format: age gender race date&time.jpg, where age is an integer from 0 to 116, gender is an integer: 0 - male, 1 - female, race is an integer: 0 - white, 1 - black, 2 - asian, 3 - indian, 4 - others, date and time, denoting when the picture was taken. The annotation was obtained from the filenames by parsing them. After that the invalid entires, such as unlabelled or partially labelled rows were removed from the dataset. Finally the images were resized and column wise 0-1 normalisation was performed on them before being used for training the gender and ethnicity classifier model.

#### 4.3.2 Software and Hardware Configuration

The experiments have been performed on an Intel Core i5-6500 3.2 Ghz processor with 4 cores. The chipset used was of the Intel Skylake Series and a RAM of 8 GB @ 3000 Mhz was used. All the experiments have been performed in Python language on the platform of Google Colaboratory.

## 5 Results

The results obtained verify that the integrated dataset performs better at ASD detection compared to the individual datasets, namely behavioural data, visual data version 1 and visual data version 9.

This result is evident across all three decision level fusion techniques, with the highest set of classification accuracies being obtained for the integrated dataset that was composed of the visual data version 1 and the behavioral data. The accuracy of the randomly integrated data where the records were combined arbitrarily was significantly less than the accuracy of the integrated dataset. The sensitivity, specificity, precision, recall and F1 scores are calculated for each of the experiments performed.

**Table 1:**
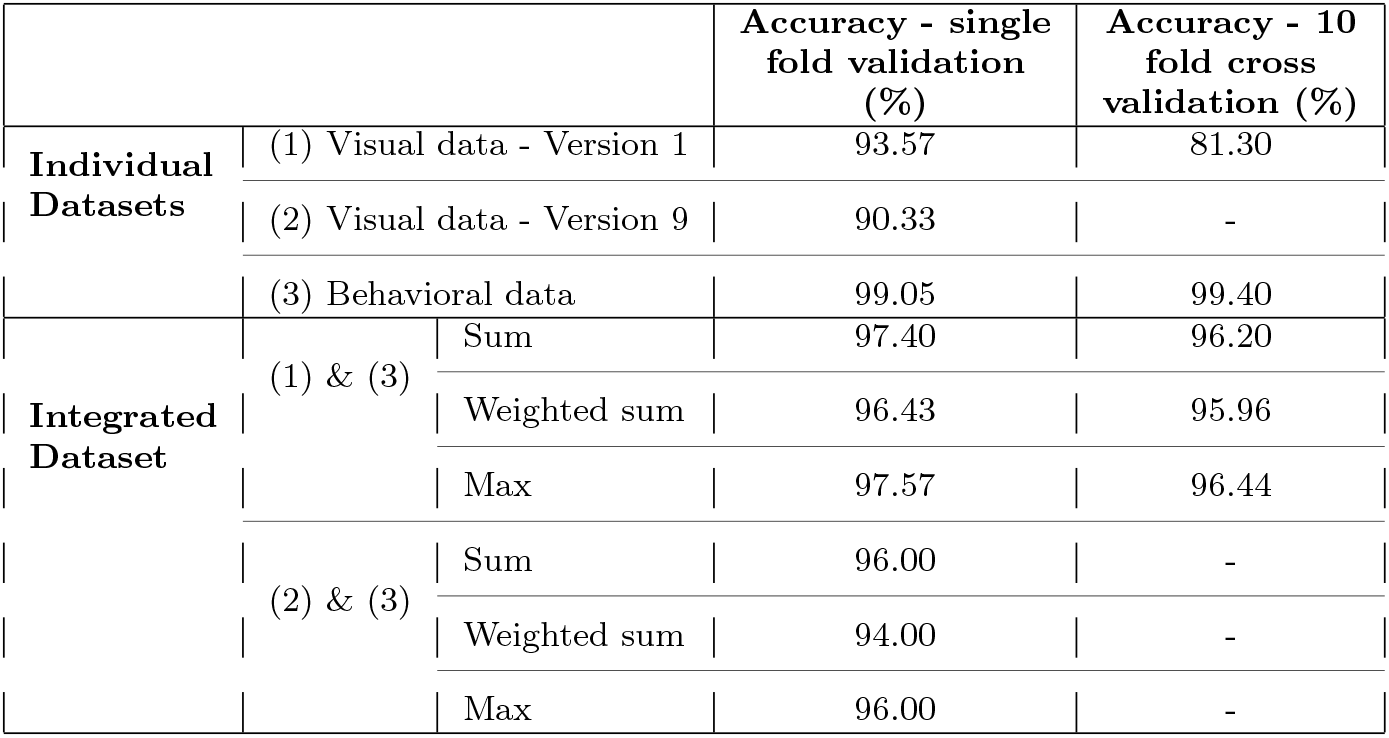
Comparison of Individual and Integrated Datasets

**Table 2:** Comparison of Integrated Dataset with Randomly Integrated Data

|  |  | Accuracy (%) |  |
| --- | --- | --- | --- |
|  |  | Visual data (Version 9) | Visual data (Version 1) |
| **Randomly Integrated Data** | Sum | 52.00 | 84.65 |
|  | Weighted Sum | 51.00 | 76.75 |
|  | Max | 55.55 | 69.64 |
| **Integrated Dataset (Grouped by common features)** | Sum | 96.00 | 97.40 |
|  | Weighted Sum | 94.00 | 96.43 |
|  | Max | 96.00 | 97.57 |

## 6 Discussion

The improved accuracy results for the experiments done with visual data version 1 are more likely to be caused due to the the presence of duplicates. For further analysis and detection of any possible overfitting, a 10 fold cross validation was performed on the visual data version 1, behavioral data and the integrated dataset formed after combining them. As can be seen from the table above, the accuracy values after 10 fold cross validation does not deviate significantly from our reference model, thus validating our model.

The accuracy of the integrated dataset created from data clusters grouped by gender and race features that are common in both datasets is significantly higher. This demonstrates the effectiveness of combining records based on common attributes. As before, the integrated dataset containing visual data version 1 has higher accuracy than the dataset containing visual data version 9 due to duplicates.

**Figure 11:**
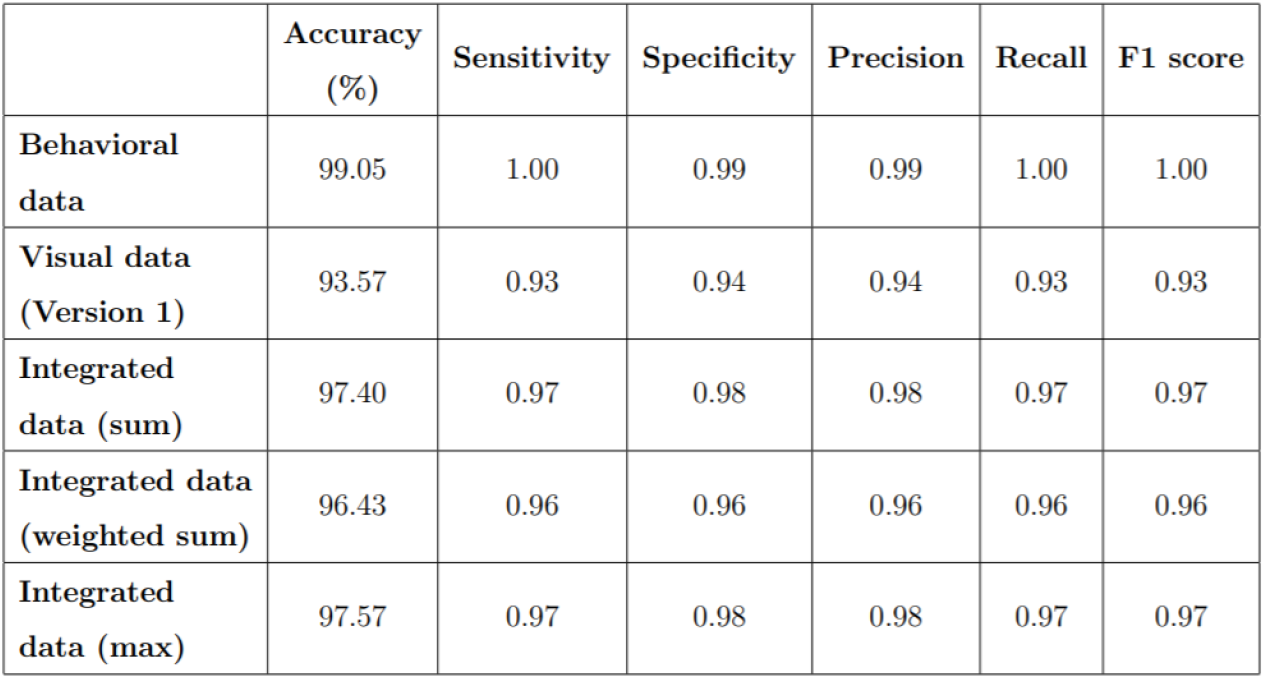
Table of Comparison of Evaluation metrics.

The results reflected from the evalutaion metrics are aligned with the assumption that integration of datasets improves the accuracy of detection of ASD. This is due to the accuracy of the integrated dataset being higher than the accuracy of visual dataset individually. The integrated dataset contains same samples present in the individual datasets, so discrepancies in the accuracy measures are more likely to be caused due to the integration.

## 7 Conclusion and Future Work

We have achieved an accuracy of 97.57% in autism detection from visual and behavioral data. Our work proves that integration of multi modal data (visual and behavioral) can be better for detecting ASD compared to individual visual dataset(93.57%). We have also shown that combining data within sub classes based on common attributes like gender and race, is more accurate than combining them arbitrarily. The datasets used in ASD studies usually consist of small number of subjects and only one mode of data. Through our work we have also developed a novel, integrated dataset which consists of multi modal data (visual and behavioral).

However, we believe the evaluation would be more accurate if ground truth of ASD labels for the data had been obtained via case studies on actual ASD patients. As shown in [27], we need to be aware of the pitfalls of machine learning when evaluating the performance of these algorithms in the ASD domain. The image data had duplicates present and there was lack of ground truth labels for the predicted image metadata. Both the datasets used for integration had skewed distributions, which may have reduced the accuracy. The dataset used for behavioral data was not robust, and using alternative datasets should give greater significant improvement to results. Finally, our prediction algorithms can only predict the race, not the ethnicity of a subject. Race and ethnicity have variations among them, race being related to the physical characteristics of the person and ethnicity being the culture that their ancestors identify with. Thus considering ethnicity and race to be the same may have introduced some inaccuracies.

For future improvements to the experiments, we can incorporate handcrafted visual features like Masculinity, Eye contact, Patterns of expression that have been shown in previous studies to be useful for ASD detection. Case studies can be performed to ascertain ground truth of the labels. So far, three simple types of Decision Level Fusion techniques have been attempted on the dataset. If the performance of the dataset is not adequate, a neural network could be used that takes the score values as input and computes the class prediction.

## Data Availability

All XXX files are available from the XXX database (accession number(s) XXX, XXX.)

## 9 Declarations

### 9.1 Consent to participate

Not Applicable.

### 9.2 Consent for publication

Not Applicable.

### 9.3 Code Availability

The integrated data and code that support the findings of this study are available upon reasonable request.

### 9.4 Authors’ contributions

All authors contributed to the study conception and design. Material preparation, data collection and analysis were performed by Nafisa Sadaf Hriti, Karishma Shaer and Farhan M Nafis Momin. The first draft of the manuscript was written by all authors and all authors commented on previous versions of the manuscript. All authors read and approved the final manuscript.

### 9.5 Compliance with Ethical Standards

This article relies on third party data that does not pose privacy risks and does not contain any studies involving human participants or animals performed by any of the authors.

### 9.8 Ethical Conduct

The study was entirely based on publicly available online datasets. For this type of study, informed consent is not required.

## References

[1] https://www.cdc.gov/ncbddd/developmentaldisabilities/facts.html, last accessed on 07/12/20

[2] Minissi, M.E., Chicchi Giglioli, I.A., Mantovani, F., Alcaniz Raya, M.: Assessment of the autism spectrum disorder based on machine learning and social visual attention: A systematic review. Journal of Autism and Developmental Disorders, 1–16 (2021)

[3] Cooper, J.O., Heron, T.E., Heward, W.L., et al.: Applied behavior analysis (2007)

[4] https://theconversation.com/science-that-could-improve-the-lives-of-people-with-autism-is-being-ignored-39951, last accessed on 26/08/21

[5] Parikh, M.N., Li, H., He, L.: Enhancing diagnosis of autism with optimized machine learning models and personal characteristic data. Frontiers in computational neuroscience 13, 9 (2019)

[6] Zunino, A., Morerio, P., Cavallo, A., Ansuini, C., Podda, J., Battaglia, F., Veneselli, E., Becchio, C., Murino, V.: Video gesture analysis for autism spectrum disorder detection. In: 2018 24th International Conference on Pattern Recognition (ICPR), pp. 3421–3426 (2018). IEEE

[7] Duan, H., Zhai, G., Min, X., Che, Z., Fang, Y., Yang, X., Gutierrez, J., Callet, P.L.: A dataset of eye movements for the children with autism spectrum disorder. In: Proceedings of the 10th ACM Multimedia Systems Conference, pp. 255–260 (2019)

[8] https://www.kaggle.com/gpiosenka/autistic-children-data-set-traintestvalidate, last accessed on 07/12/20

[9] Helland, W.A., Helland, T.: Emotional and behavioural needs in children with specific language impairment and in children with autism spectrum disorder: The importance of pragmatic language impairment. Research in Developmental Disabilities 70, 33–39 (2017)

[10] Andrews, D.S., Marquand, A., Ecker, C., McAlonan, G.: Using pattern classification to identify brain imaging markers in autism spectrum disorder. In: Biomarkers in Psychiatry, pp. 413–436. Springer, ??? (2018)

[11] Mujeeb Rahman, K., Monica Subashini, M.: A deep neural networkbased model for screening autism spectrum disorder using the quantitative checklist for autism in toddlers (qchat). Journal of Autism and Developmental Disorders, 1–15 (2021)

[12] Tan, D.W., Gilani, S.Z., Maybery, M.T., Mian, A., Hunt, A., Walters, M., Whitehouse, A.J.: Hypermasculinised facial morphology in boys and girls with autism spectrum disorder and its association with symptomatology. Scientific reports 7(1), 1–11 (2017)

[13] Beary, M., Hadsell, A., Messersmith, R., Hosseini, M.-P.: Diagnosis of autism in children using facial analysis and deep learning. arXiv preprint 2008.02890 (2020)

[14] https://www.kaggle.com/fabdelja/autism-screening-for-toddlers/version/1?select=Toddler+Autism+dataset+July+2018.cs, last accessed on 07/12/20

[15] Akter, T., Satu, M.S., Khan, M.I., Ali, M.H., Uddin, S., Lio, P., Quinn, J.M., Moni, M.A.: Machine learning-based models for early stage detection of autism spectrum disorders. IEEE Access 7, 166509–166527 (2019)

[16] http://fcon1000.projects.nitrc.org/indi/abide/, last accessed on 07/12/20

[17] Thabtah, F., Peebles, D.: A new machine learning model based on induction of rules for autism detection. Health informatics journal 26(1), 264–286 (2020)

[18] Niu, K., Guo, J., Pan, Y., Gao, X., Peng, X., Li, N., Li, H.: Multichannel deep attention neural networks for the classification of autism spectrum disorder using neuroimaging and personal characteristic data. Complexity 2020 (2020)

[19] Jung, S.-G., An, J., Kwak, H., Salminen, J., Jansen, B.J.: Assessing the accuracy of four popular face recognition tools for inferring gender, age, and race (2018)

[20] Chandaliya, P., Kumar, V., Harjani, M., Nain, N.: SCDAE: Ethnicity and Gender Alteration on CLF and UTKFace Dataset, pp. 294–306 (2020). https://doi.org/10.1007/978-981-15-4018-927

[21] Kärkkäinen, K., Joo, J.: Fairface: Face attribute dataset for balanced race, gender, and age. arXiv preprint arXiv:1908.04913 (2019)

[22] Li, Y., Ngom, A.: Data integration in machine learning. In: 2015 IEEE International Conference on Bioinformatics and Biomedicine (BIBM), pp. 1665–1671 (2015). IEEE

[23] Gokberk, B., Akarun, L.: Comparative analysis of decision-level fusion algorithms for 3d face recognition. In: 18th International Conference on Pattern Recognition (ICPR’06), vol. 3, pp. 1018–1021 (2006). IEEE

[24] Kim, J.I., Bang, S., Yang, J.-J., Kwon, H., Jang, S., Roh, S., Kim, S.H., Kim, M.J., Lee, H.J., Lee, J.-M., et al.: Classification of preschoolers with low-functioning autism spectrum disorder using multimodal mri data. Journal of Autism and Developmental Disorders, 1–13 (2022)

[25] Casa, A., Bouveyron, C., Erosheva, E., Menardi, G.: Co-clustering of time-dependent data via the shape invariant model. Journal of Classification 38(3), 626–649 (2021)

[26] http://aicip.eecs.utk.edu/wiki/UTKFace, last accessed on 02/03/20

[27] Bone, D., Goodwin, M.S., Black, M.P., Lee, C.-C., Audhkhasi, K., Narayanan, S.: Applying machine learning to facilitate autism diagnostics: pitfalls and promises. Journal of autism and developmental disorders 45(5), 1121–1136 (2015)

